# Bayesian network approach to develop generalisable predictive model for COVID-19 vaccine uptake

**DOI:** 10.1101/2023.01.31.23285300

**Authors:** Raghav Awasthi, Aditya Nagori, Bouchra Nasri

## Abstract

The effectiveness of a vaccine depends on vaccine uptake, which is influenced by various factors, including vaccine hesitancy. Vaccine hesitancy is a complex socio-behavioral issue, influenced by misinformation, distrust in healthcare providers and government organizations, fear of side effects, and cultural or religious beliefs. To address this problem, AI models have been developed, but their global generalizability remains unclear. Therefore, this study aimed to identify global determinants of vaccine uptake and develop a generalizable machine learning model to predict individual-level vaccine uptake. The study used publicly available survey data from 23 countries and employed Bayesian networks and generalized mixed effects models to identify key determinants of vaccine uptake. The results showed that trust in the central government and vaccination restrictions for national and international travel were key determinants of vaccine uptake. A generalized mixed effects model achieved an AUC of 89% (SD=1%), precision of 90% (SD = 4%), and recall of 82% (SD=2%) on unseen testing data from new countries, demonstrating the model’s generalizability. The findings of this study can inform targeted interventions to improve vaccine uptake globally.

## Introduction

Vaccination is a crucial public health measure that helps to prevent the spread of infectious diseases and protect individuals and communities [1]. However, the success of vaccination programs depends on the willingness and ability of individuals to receive vaccines [2]. Formally, Vaccine hesitancy according to the *Strategic Advisory Group of Experts on Immunization (SAGE)* Working Group [2,3] is defined as a delay in accepting or refusing vaccination [4,5]. Upon the release of COVID-19 vaccines, it was essential that a substantial percentage of the population need to get vaccinated [4] [7] [6] to achieve herd immunity. For the decision-makers it becomes crucial to comprehend and recognise a person’s vaccination-related behavior. The vaccination hesitation behavior of an individual is influenced by variables such as vaccine accessibility, geographical location, cost, socio-economic factors, quality of healthcare, and most importantly, trust in vaccines and health experts [7,8]. Artificial intelligence (AI) techniques, which involve processing large amounts of data, identifying patterns and trends, could be helpful in making predictions that might guide the understanding and the implementation of efficient vaccination strategies. Several researches have shown that AI-based techniques may be used to identify important vaccination uptake factors and predict vaccine uptake behavior at the individual level [9–13]. However, models developed in previous literature [8–12] are based on a local scale (for a specific region or country), and none of them studied whether or not vaccination policies can be implemented on a global scale. The success of vaccination programmes may vary between geographical location and socio-demographic factors of a specific population [7]. The capacity of AI algorithms to effectively predict the effects of vaccination programmes in diverse countries and people may be impacted by factors such as cultural, economic, and political disparities [12,14–16]. Additionally, there may be variations in data availability and quality, as well as in the knowledge and resources available to create and deploy AI algorithms, among locations and populations[16,17].

In this study, we aim to analyze the global generalizability of AI-driven vaccination policies, with the focus of identifying the key determinants of vaccine uptake behavior and developing a generalizable machine learning model to predict the vaccine uptake behavior at the individual level. The findings of this study will provide valuable insights into the global determinants of COVID-19 vaccine hesitancy and the generalizability of prediction models for COVID-19 vaccine hesitancy. These insights will be useful for policymakers, public health officials, and other stakeholders in their efforts to address COVID-19 vaccine hesitancy and promote vaccination uptake. Finally, as part of this study, a AI-driven tool is developed to help researchers and policymakers in analyzing the vaccine uptake behavior based on survey data for a specific disease or one/multiple geographical locations.

## Results

### Data characteristics

The descriptive statistical analysis showed some pattern of vaccine hesitancy on socioeconomic factors, opinions on COVID-19 and COVID-19 vaccine, trust on government, doctors and employer, and necessity of COVID-19 vaccine on college, school, travel and employment. A descriptive association between behavioral factors, socioeconomic factors and vaccine hesitancy and key determinants of policy is given in **Figure 1**. We found that the proportion of respondents who are hesitant to receive vaccines decreases with age. Similarly, respondents with higher anxiety disorders tend to have a lower probability of vaccine hesitancy. Change in household income is a strong indicator of behavior change. In fact, we found that people whose household income is severely affected during COVID-19 are less likely to have vaccine hesitancy. Overall data depicts the disparities in vaccine hesitancy among people of various ages, mental health conditions, trust-related behavior indicators, and impact of COVID-19 related indicators. Based on the prevalence of vaccination hesitancy across countries, we found that Russia has the highest rate (11.51%), followed by Poland (8.23%) and South Africa (7.95%). Moreover, we found that India has the lowest rate of vaccination hesitancy, at 0.57%.

**Figure 1:**
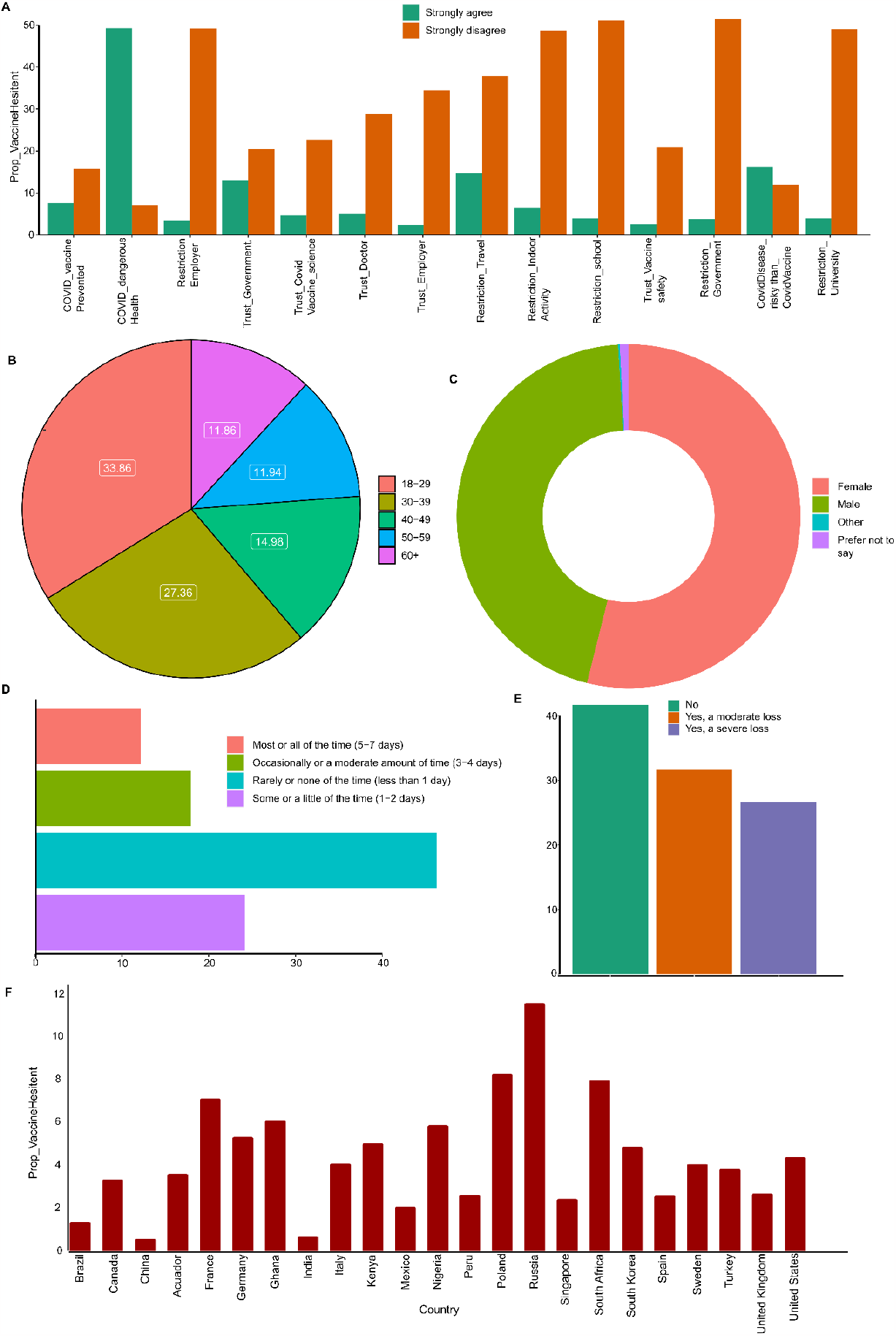
**A)** Proportion of vaccine hesitancy characterized based on opinions. **B)** Distribution of vaccine hesitancy proportion on different age-groups. **C)** Distribution of vaccine hesitancy proportion on different genders. **D)** Vaccine hesitancy and recent trends of the number of days that a respondent felt depressed. **E)** Vaccine hesitancy and household income loss during COVID-19 **F)** Vaccine Hesitancy distribution by Country.

### Bayesian Network structure learning and network community/cluster detection to visualize global determinants

In the learned bootstrapped network (**Figure 2B**), we have only considered the nodes’ connections that have edge strengths greater than 0.51(majority of voting in bootstrapped network). Then, in the bootstrapped Bayesian network, we first performed node clustering to visualize the set of key indicators that have a high impact on the outcome variable. Our algorithm detected 11 clusters of nodes, and the largest cluster contains 15 nodes (**Figure 2A, Figure 3A**). Our outcome variable was found in the 5 clusters (Clusters # 2,3,4,7,8) among all 11 identified clusters. Every cluster includes some specific features. For example, cluster #1 compromise about socio-demographic information such as age, gender, and monthly income. Similarly, cluster#8 has 15 nodes, making it the largest cluster. This cluster includes variables associated to COVID-19 and COVID-19 vaccinations, and household income changes during COVID-19. With trust in stakeholders (Doctor, Employer, Government), COVID-19 vaccination necessity for international travel, and indoor activities. Since communities reveal the clusters of variables, it is also necessary, from a policy perspective, to determine which variables are mainly affecting vaccine hesitancy. Next, we calculated the markov blanket of outcome variables in order to derive conclusions about the main factors influencing our outcome variable.

**Figure 2:**
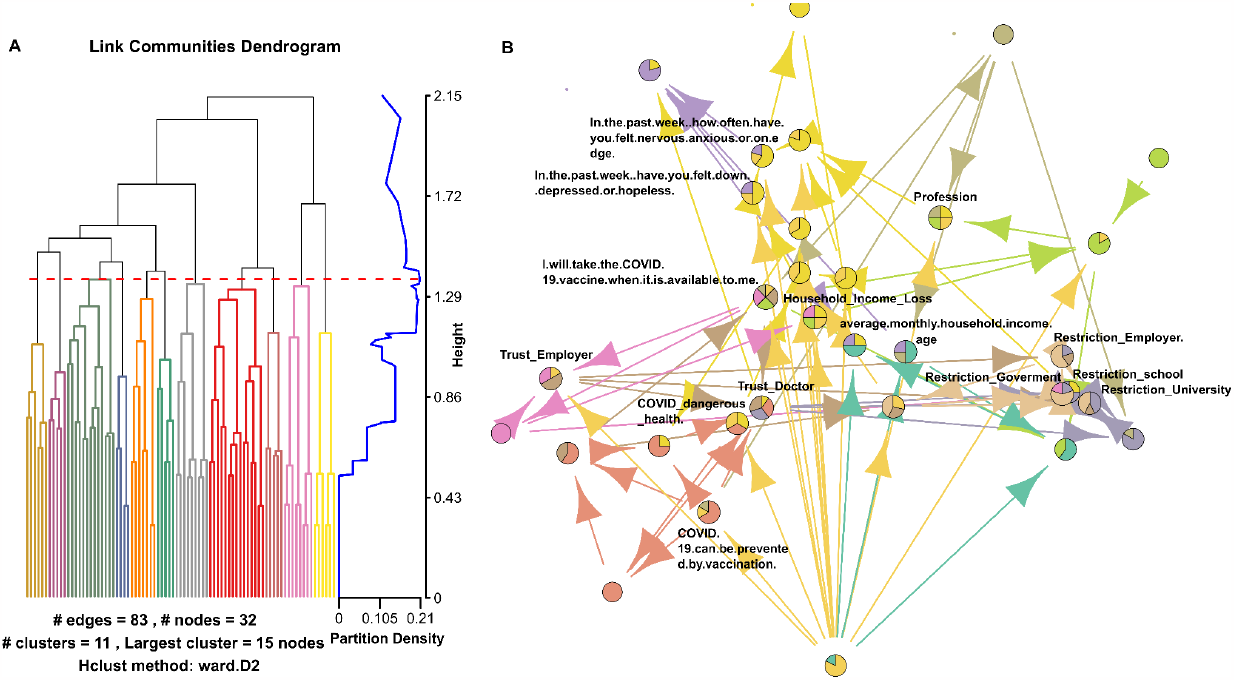
**A) Extracting link communities from the entire Bayesian network:** 11 clusters/communities were identified in the Bayesian network with the largest cluster of 15 nodes. **B) Network Structure with learned community:** To aid the visualization of key nodes we limit the display of nodes that belong to 3 or more communities.

**Figure 3:**
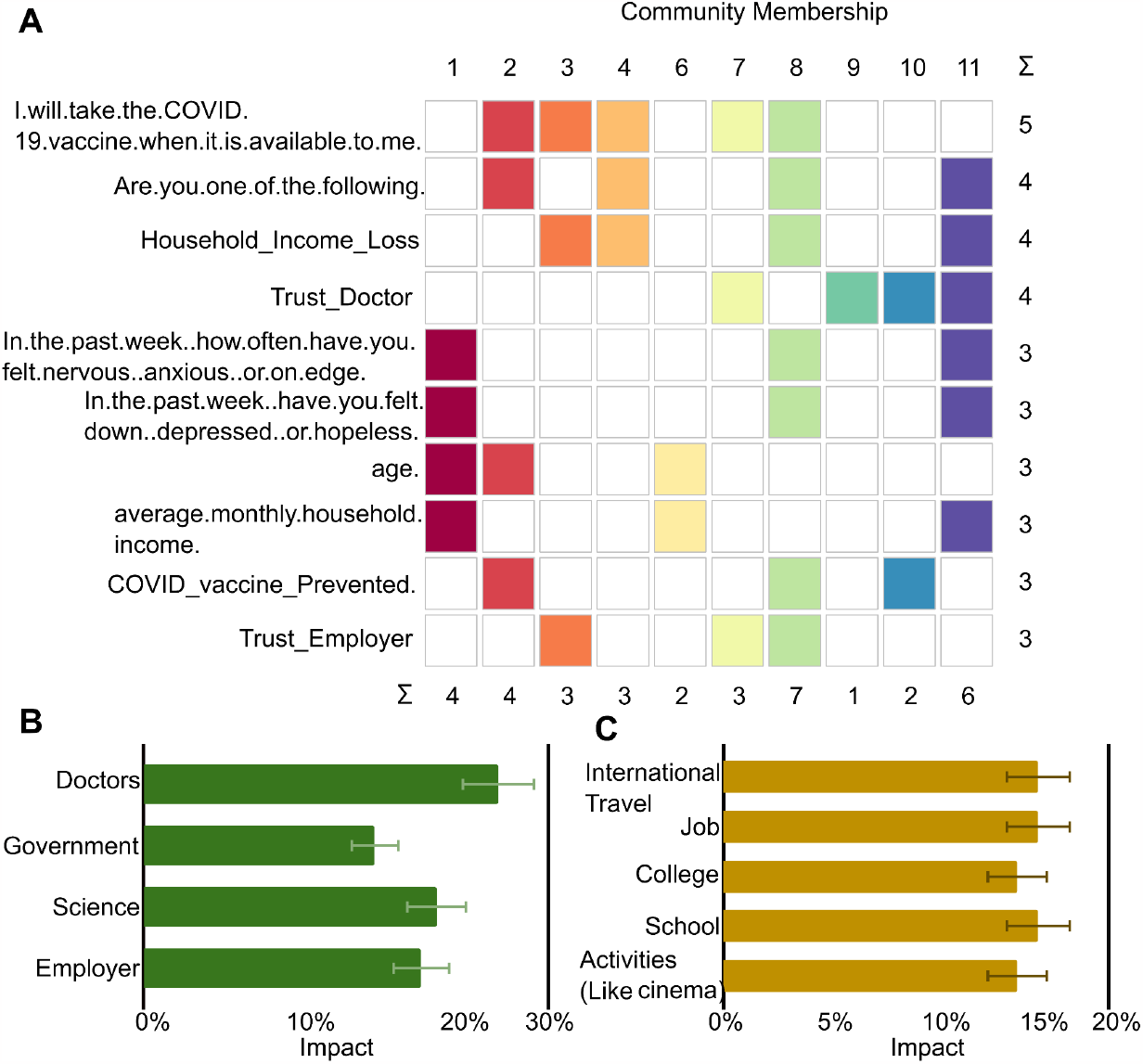
A) Node community membership for the top-connected nodes: B) Association of vaccine uptake with trust on stakeholders C) Association of vaccine uptake with public activity.

From the markov blanket analysis, we found that trust and restriction are the major categories of intervention that impact vaccine hesitancy. Furthermore, our analysis will be focused on these two categories.

### Association between trust towards stakeholders and vaccine uptake

The results showed that the public’s trust in COVID-19 vaccines and vaccination is a key for vaccines’ success **(Figure 3B)**. Our Bayesian network analysis found that people’s faith in their doctors, government officials, and the science behind COVID-19 had a substantial effect on their vaccination behavior. Our analysis revealed a 23% increase in the likelihood that a person would get a vaccination if they had confidence in physicians. Similarly, we discovered a 23% increase in the likelihood of vaccination acceptance when individuals had confidence in government authorities. The Bayesian network also identified confidence in the science behind COVID-19 as a vital element for vaccine uptake behavior, and a 19% increase in the likelihood of vaccine uptake behavior if individuals had faith in the research underlying COVID-19. Also, the results have shown that a person’s likelihood of getting a vaccination goes up by 18% if they trust their employer.

### Association between social restrictions and vaccine hesitancy

COVID-19 has halted the socio-economic vehicles of the entire world, which was required to lessen the acceleration of viral spread. Multiple sectors, including travel, employment, colleges, social gatherings, and events, have mandated comprehensive COVID-19 immunization since its introduction. This mandate has raised the vaccination rate among individuals. We found that foreign travel limitations have boosted the likelihood of vaccination uptake by 15%. Mandatory vaccines in workspace and in schools/universities (colleges) limits in job colleges and schools have raised the likelihood of vaccination uptake by 15% and 14%, respectively. With the advent of cinema and concerts, it is believed that these activities would become the focal point of social gatherings and in certain countries, vaccination were also made obligatory. We found that vaccination requirement criteria had led to a 14% rise in the decision to get vaccinated **(Figure 3C)**.

### Prediction of vulnerable individuals to maximize vaccine uptake

We found based on the GLMM that the Trust on the central governments as the one the key predictors with log-odds ratio greater than 0.55[0.25, 0.84](p-value< 0.0002), is equally significant across different income groups as shown in **Figure 4B**. We also found that the restrictions on international travel as important trigger for vaccine uptake with log-odds of 0.4 [0.15, 0.65](p-value= 0.0034) for higher income countries, 0.64 [0.35, 0.93](p-value= 0.00016) for lower middle income countries, and 0.79 [0.54, 1.04] (p-value= 3.69*10^−10^) for upper middle income countries. Age group-wise, individuals with age-group 60+ have higher positive odds of vaccine uptake in lower middle income countries contrary to higher and upper middle income countries **(Figure 4A, 4B)**. Income groups having higher than the country median income have significant odds of vaccine uptake.

**Figure 4:**
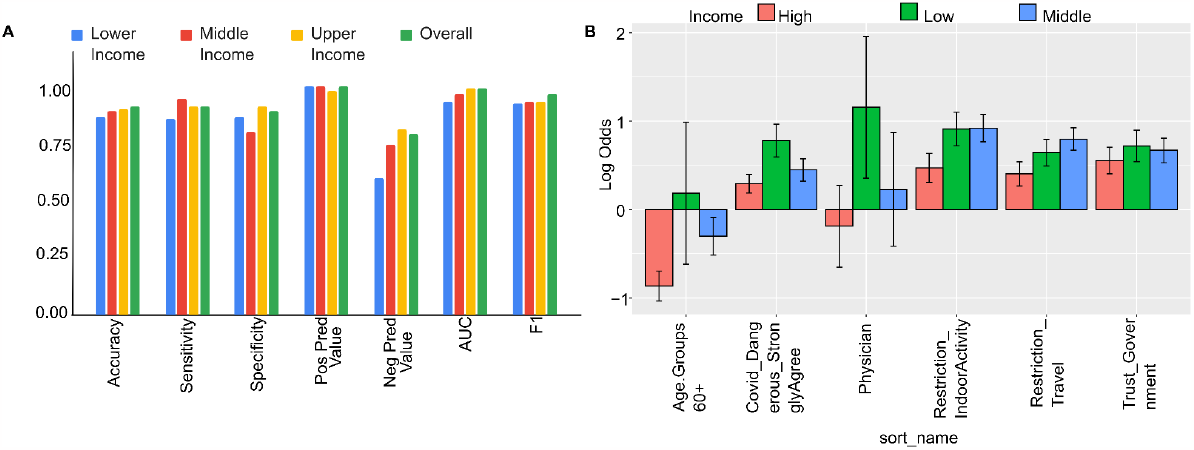
**A) Prediction model results :** Modeling results using Generalized mixed effects models, model achieved an AUC of 89% for the generalized prediction across different countries with Positive predictive value of ∼90% with Sensitivity of 82%. **B) Top 5 features in prediction**: The top 5 features were selected from the Mixed effects model based on their coefficient magnitude in the model for low, middle, and high income countries. We plotted the coefficient estimate (Log odds) of the top features’ union (Low Income, Middle Income, and High Income), and the error bar represents the standard error. In the prediction model, we again find the trust and social restriction indicators to be the most important.

## Discussion

Multiple stakeholders, including government decision-makers, community, health personnel, media sources, and internet platforms, impact vaccination rates [18]. These factors may encourage or discourage vaccination by fostering more or less permissive environments. Consequently, it is essential to evaluate how the health system factors may impact public behavior. Several data-driven models and theoretical studies have been done to uncover vaccine uptake indicators. Other machine-learning models were also built to predict the vaccine adherence paradigm [19–22].

Here, we analyzed data from various countries to determine the variables influencing vaccination rates. We also evaluated the performance of machine learning prediction models in high-income, lower-middle-income, and high-middle-income nations. We found that our models outperformed similar efforts in the literature with a high margin of accuracy compared to studies in the literature [35-37]; however, there were few differences concerning the choice of data and methods. Previous studies like Lincoln et al surveyed individuals from five different high income countries to understand factors driving vaccine hesitancy during the COVID-19 pandemic. Results showed that only 57.4% of participants indicated they would definitely or probably get vaccinated. A machine learning model was used to identify predictors of vaccine hesitancy, such as vaccination conspiracy beliefs, mistrust, age, income, and population density, with accuracy (82% sensitivity and 79-82% specificity) [23]. Carrieri et al. used the supervised random-forest machine-learning algorithm on area-level indicators of institutional and socioeconomic backgrounds to predict the vaccine hesitancy rate for Italian local authorities, which helped public-health practitioners run targeted awareness campaigns. The study found that non-clinical features like employment rate, proportion of waste recycling had the highest predictive powers in the random-forest algorithm, with an AUC of 0.836 [24]. Santillana et al. used Google searches, Twitter microblogs, nearly real-time hospital visit records, and data from a participatory surveillance system data with ensemble methods of stacking regression that combine separate outcomes from each model of different statistical classifiers to predict vaccinations and achieved a testing accuracy of 85.71% in the 10-fold cross-validation [25]. This study examines the state-level features and policies that are most important in achieving a threshold level vaccination rate using a decision tree algorithm on a dataset of all states in the US. The study finds that workplace travel is the most important predictor of vaccination rates. The study also uses alternative algorithms to confirm these findings, with accuracy ranging from 80-88% and sensitivity from 92.5-100%. The findings provide actionable policy insights to increase vaccination rates and combat the COVID-19 pandemic [26].

However, previous studies have focused on specific geographical areas and have not investigated the global factors that influence vaccination rates or the generalizability of machine learning models for predicting vaccine hesitancy. In this study, using a survey questionnaire with 23135 respondents across 23 countries. We used Bayesian networks and generalized linear models to identify the critical determinants of vaccine uptake. We then used Markov Blankets obtained from the Bayesian networks to estimate the vital predictors of vaccine uptake, which were used to build our models. We tested the generalizability of our models across countries and income groups. Our study found that trust in doctors had a greater impact on vaccination behavior than trust in other stakeholders. Factors such as faith in vaccine efficacy and safety, and trust in institutions and government decisions also play a role in vaccination uptake. The use of social constraints, such as restrictions on certain activities for those who are not vaccinated, may also influence behavior [39-40]. Further, our generalized mixed effects model approach achieved an AUC of 89%, Precision of 90%, and Recall of 82% on the prediction task on new countries, indicating its potential effectiveness in predicting vaccine uptake in contemporary populations.

Our study has several strengths, including large sample size and a rigorous methodology for identifying and predicting vaccine uptake behavior. Our approach also has the potential to be applied on a global scale, making it a valuable tool for policymakers looking to improve vaccination programs. However, our study has some limitations, like the fact that it is challenging to derive causal influences from observational data, and our study does not claim to do that. However, the usefulness of Bayesian networks as probabilistic reasoning systems is well known but little applied in public health settings, including we believe that additional data is required to explore in more detail and determine the feasibility of AI-driven techniques for enhancing global vaccination programmes. In addition, since the study deals with sensitive public health issues, the responses to the survey questions may have some degree of erroneousness. Hence, it is difficult to corroborate the associations that were discovered in a data-driven way.

Finally, as a conclusion, despite the development of numerous machine learning models in public health, there are only a handful of models that are explainable and generalizable. In this study, we provided a framework that not only identified explainable policy solutions for COVID-19 vaccination but could also be adopted for any data-driven interventional modeling studies.

## Data and Methods

### Data

We have utilized publicly available survey data for our analysis [27]. Data consists of 23,135 participants from 23 countries who participated in the survey. About half (50.2%) were female and resided in LMICs (Low and Middle income Countries) (52.2%), three-fifths (59.9%) were between the ages of 30 and 59, and one-fifth (22.4%) were college graduates. The data represents 23 countries: Brazil, Canada, China, Ecuador, France, Germany, Ghana, India, Italy, Kenya, Mexico, Nigeria, Peru, Poland, Russia, Singapore, South Africa, South Korea, Spain, Sweden, Turkey, the UK, and the US. The survey questionnaire was made by a panel of experts based on COVID-19 vaccine acceptance studies as well as studies on pandemic control measures and vaccination intentions [28,29]. The questionnaire is composed of 31 questions (***See S*upplementary information of** [27]). These questions underline mainly the three following concepts: 1) Questions representing perceptions of risk (q1) (q3), efficacy (q2), safety (q4), and trust (q5 and q6) as important individual determinants of COVID-19 vaccine hesitancy and of routine immunization; 2) questions regarding the status of vaccine regarding the first dose (q7) and the vaccine hesitancy which based on taking or no vaccine when available(q8) 3) questions related to socio-economic factors like education (q29), income (q30), and geographical location (q31).

### Methods

To begin our analysis, we utilized a probabilistic graphical model that seeks to identify key factors underlying vaccine hesitancy. Then using a supervised machine learning approach, we have examined the generalisability of vaccination uptake prediction models across countries. **Figure 5** summaries the steps used in the analysis. First we cleaned and preprocessed the data-in order to handle missing information. Our outcome variable regarding vaccine hesitancy corresponds to answers related to q8 of the survey which is *‘I will take the COVID-19 vaccine when it is available to me*’. We trained a bootstrapped Bayesian network in order to quantify the interaction between data variables and learn the determinants of global vaccination hesitancy. Using the learnt Bayesian network, we computed Markov blanket variables for outcome variables. Further Markov blankets were used to predict the outcome variable. Finally, we calculated the generalizability of the prediction model across countries.

**Figure 5:**
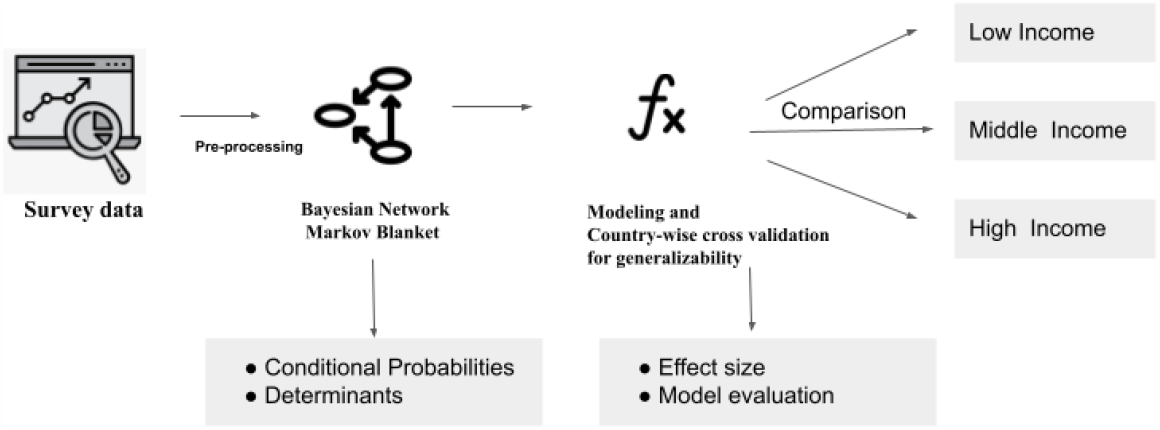
Summary figure of the overall analysis, survey data was pre-processed to find the determinants of vaccine uptake using bayesian network markov blanket approach, conditional probabilities were analyzed for inference. The important determinants were used to build generalizable models and cross-validated on new countries, finally the effect sizes of determinants were computed and analyzed with income group stratification.

### Bayesian Network Analysis for Learning Global Determinants of Vaccine Hesitancy

A Bayesian network [30] is a probabilistic graphical model that depicts the joint probability distribution of a group of random variables with the potential for mutual causation. The Bayesian network is a directed acyclic graph with nodes representing random variables, edges between pairs of nodes expressing the causal link between these nodes, and a conditional probability distribution inside each node. The primary objective of Bayesian network analysis is to model complex systems and understand the relationships between different variables [31,32]. In our study, a Bayesian network model will be used to identify the key determinants of vaccine hesitancy and understand how these determinants interact with each other. We used a robust structure of the learnt Bayesian network (BN) by including a bootstrapping and ensemble averaging of edge directions in order to overcome the complex dependencies with potential confounding effects, mediation, and intercausal dependency regarding our outcome [33–36]. The best probabilistic graphical model that adequately explained the data was chosen using the hill climbing optimizer and a score based on the Akaike information criteria [37]. Over 11 BNs, we used bootstrapped learning with majority voting to learn Bayesian networks [32]. The package *wiseR* in R was used to do the analysis [38]. Furthermore, for community (clusters among nodes in BN) detection in the learnt network, we have used *linkcomm* R package[39].

### Predictive modeling

Predictive vaccine uptake behavior might be a helpful tool for public health professionals, as it can help identify populations at risk of vaccine hesitancy and allow targeted interventions to be implemented in order to increase vaccine uptake. We used the GLMM to predict vaccine uptake among the survey respondents [40,41]. Variable Country was used as the random effect. We used the variables obtained in the markov blanket bayesian network analysis as the important predictors. The model equation is given by

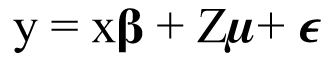

where, y is nx1 matrix of binary responses, i.e. vaccine uptake agree or disagree, x is nxp matrix of predictors, important survey questionnaire as obtained from the markov blanket, **β** is the p x1 vector of regression coefficients, Z is the Nxq for q random effects, i.e. Country. ***μ*** is qx1 vector of random effects and ***ϵ*** is a Nx1 vector of residuals that of y is not explained by model.

For testing the performance of models on new countries, we divided data into country-wise 5 fold splits, with 80% countries in the model development and 20% countries in the testing set in each split. The model evaluation resulted in generalized performance in the new countries. We also developed the income specific models and computed the effects sizes and model performances.

## Funding

This work was supported by Pathcheck foundation’s data informatics center for Epidemiology (DICE).

## Acknowledgement

We acknowledge Graham Dodge, president at PathCheck foundation and Bethany LoMonaco, Director, strategy and development for their constant support.

## Data availability statement

Data used in this study is publicly available and easily downloadable under the “*Source Data*” section of the article at https://www.nature.com/articles/s41467-022-31441-x. Data is also available at the public repository https://doi.org/10.5281/zenodo.6560427.

## Notes

### Competing Interest Statement

The authors have declared no competing interest.

### Summary of Updates

Made correction with the language.

